# National Trends and Geographic Disparities in Ischemic Heart Disease Mortality in the United States, 1968–2023

**DOI:** 10.1101/2025.11.05.25339633

**Authors:** Muhammad Umar, FNU Kritika, Sahil Jairamani, Diya Rathi, Mirza Mohammad Ali Baig, Anchit Chauhan, Eugene Yang, Avirup Guha, Harikrishnan Hyma Kunhiraman, Jamal S. Rana, Alrumaisa Alhassan

## Abstract

**Background:** Ischemic heart disease (IHD) is a leading cause of mortality in the United States, with significant variations across demographic and geographic factors. This study analyzes trends in IHD-related mortality among adults (> 25 years) from 1968 to 2023 using the CDC WONDER database.

**Methods:** We analyzed death certificates of adults aged >25 years from the CDC-WONDER database with IHD (ICD-8 codes: 410-413, ICD-9 codes: 410-414 and ICD-10 codes: I20-I25) from 1968-2023. Age-adjusted mortality rates (AAMR) per 100,000 population were stratified by gender, race, census region, and year. Join-Point analysis was performed to estimate annual percent change (APC) and average annual percent change (AAPC), complemented by spatiotemporal modeling to capture the geographic variations and evolving patterns of IHD mortality over time.

**Results:** Between 1968 and 2023, IHD caused 27,685,173 deaths in adults aged >25 years. Throughout the study period, the overall AAMR demonstrated a sustained and significant decline, falling from 746.29 in 1968 to 127.07 in 2023 with an AAPC (−3.12; 95% CI: −3.51 to −2.73; p<0.001). Additionally, the overall AAMR for males (452.99) was higher than that of females (260.08). Non-Hispanic (NH) Black or African American displayed slightly higher AAMR (346.75) as compared to NH white (345.38). The Northeast had the highest overall AAMR (379.61), followed by the Midwest (351.31), the South (329.15), and the West (303.15). Older adults (≥65 years) had the highest overall crude mortality rate (CMR) (1312.66), followed by middle-aged adults (45–64 years) (171.67). Younger adults (25–44 years) had the lowest rate (12.18).

**Conclusion:** Mortality rates due to IHD have decreased in the United States over the last few decades. However, the rate of decline has decreased in the past decade. In addition, significant disparities still exist across different sexes, age, race/ethnicity, and geographic regions. These disparities highlight the need for targeted interventions and improvement of clinical care facilities in high-risk populations.

## Introduction

The epidemiological landscape of the ischemic heart disease (IHD) related mortality in the United States established a complex and evolving challenge within contemporary public health.^**1**^ Recent growth in the prevention and treatment of IHD highlights the substantial advancements in cardiovascular medicine encompassing both novel pharmacological therapies as well as the cutting edge technological innovations.^**2-4**^ These advancements in the prevention and management of IHD, along with a deeper understanding and control of its contributing factors have led to significant improvements in the patient outcomes. These developments have resulted in a significant extension of life expectancy and a substantial reduction in the mortality rates associated with the IHD.^**5**^ Yet, despite these achievements, IHD continues to enforce a major public healthcare burden and declines in the mortality rate have not been uniformly distributed across the population.^**5,6**^

Persistent disparities by sex, race, and geography highlights the need for precision public health strategies and equitable implementation of evidence based cardiovascular care.^**7,8**^These sociodemographic factors influence not only the prevalence and clinical outcomes of IHD but also access to timely diagnosis and high quality treatment. Structural inequities in the healthcare delivery contribute to uneven declines in IHD mortality highlighting the urgent need for the targeted, equity focused interventions.^**9-11**^ This study significantly advances the field by integrating the high-resolution spatiotemporal trend analysis and multilayered risk stratification which offers an unprecedentedly detailed understanding of the geographic and demographic disparities in IHD mortality. Analyzing IHD mortality trends over the time is essential for informing evidence based interventions and shaping future cardiovascular care strategies. By identifying the multifaceted factors influencing these deaths, this study aims to examine long-term trends in ischemic heart disease mortality and provide actionable insights for promoting equitable health outcomes and guiding targeted public health and clinical interventions.

## METHODOLOGY

### Study setting and population

Data on the ischemic heart disease (IHD) related mortality in the United States from the year 1968 to 2023 was obtained from the Centers for Disease Control and Prevention (CDC) Wide-Ranging Online Data for Epidemiologic Research (WONDER) database which systematically gathered the national mortality records derived from death certificates maintained by the state vital registration systems.^**12**^ We used the Multiple Cause of Death and Underlying Cause of Death public use files to identify all deaths where IHD was recorded as the primary cause of death. To maintain consistency across the study period, deaths were identified using the International Classification of Diseases (ICD) codes corresponding to the IHD for each revision period as ICD Eighth Revision, Clinical Modification (ICD-8-CM; codes 410-413) for 1968-1978, ICD Ninth Revision, Clinical Modification (ICD-9-CM; codes 410-414) for 1979-1998 and ICD Tenth Revision, Clinical Modification (ICD-10-CM; codes I20-I25) for 1999-2023. The CDC WONDER database is a publicly available, de-identified dataset and therefore the present analysis was not required for the institutional review board (IRB) approval. The study adhered to the Strengthening the Reporting of Observational Studies in Epidemiology (STROBE) reporting guidelines.^**13**^

### Data extraction

From the CDC WONDER database, we extracted the information on year of death, sex, age group, race, and region for all the deaths in which IHD was recorded as the underlying cause of death. The study included the adults aged ≥25 years in the study and were categorized into 10 year intervals to ensure the comparability across decades. Racial groups included non-Hispanic (NH) White, NH Black or African American, NH Asian or Pacific Islander, NH American Indian or Alaska Native and Hispanic or Latino. Geographical regions were classified as the Northeast, Midwest, South and West according to the U.S. Census Bureau classification.

### Data synthesis and statistical analysis

Crude mortality rates (CMRs) for IHD were calculated per 100,000 people by dividing the number of deaths each year by the U.S. population, and age-adjusted mortality rates (AAMRs) were standardized to the 2000 U.S. population to allow the unbiased comparisons over time and between different groups.^**14**^ Temporal trends in the AAMRs were examined using the Joinpoint Regression Program (v5.0.2, NCI, Bethesda, MD), which fits log linear regression models to detect the points where trends were changed significantly.^**4**^ To assess both the magnitude as well as the direction of trends in IHD related mortality the annual percentage change (APC) and average annual percentage change (AAPC) with their 95% confidence intervals were calculated. Statistical significance was assessed using the Monte Carlo permutation test and two-tailed t-tests.^**15,16**^ A significance level of p ≤ 0.05 was considered as significant. This allowed us to identify the key inflection points in the trends of mortality that capture changes in population health, risk factors, and the impact of healthcare changes over time.

### Statistical Analysis

We assessed spatial clustering of ischemic heart disease (IHD) mortality across U.S. counties from 1968 through 2020 using age-adjusted mortality rates obtained from the CDC WONDER Underlying Cause of Death database. Rates were standardized per 100,000 population.

Spatial autocorrelation was evaluated using the Getis-Ord Gi* statistic to identify significant local clusters (hotspots and coldspots) of IHD mortality. A queen contiguity spatial weights matrix was employed to define neighborhood relationships among counties, and 999 Monte Carlo permutations were conducted to determine the statistical significance of clustering. States were classified as hotspots or coldspots at 90%, 95%, and 99% confidence levels, with positive Gi* Z-scores indicating hotspots and negative Z-scores indicating coldspots.

To assess spatiotemporal dynamics, analyses were stratified into five intervals: 1968–1978, 1979–1990, 1991–1998, 1999–2010, and 2011–2020. Data from 1968 to 1978 were initially extracted but excluded from analysis because IHD-related deaths during that period were too few for meaningful spatial evaluation. For rest of the periods, we summarized the proportion of significant hotspots and coldspots, mean Gi* Z-scores, and mean IHD mortality rates. All spatial analyses were conducted using ArcGIS Pro (Esri, Redlands, CA) and R version 4.5.1 with the spdep package.

## Results

### Annual Trends

From 1968 to 2023, the age-adjusted mortality rate (AAMR) for IHD showed a substantial decline–from 746.29 to 127.07 deaths per 100,000 **(Supplementary Tables 1–3, Fig 2)**.

**Figure 1:**
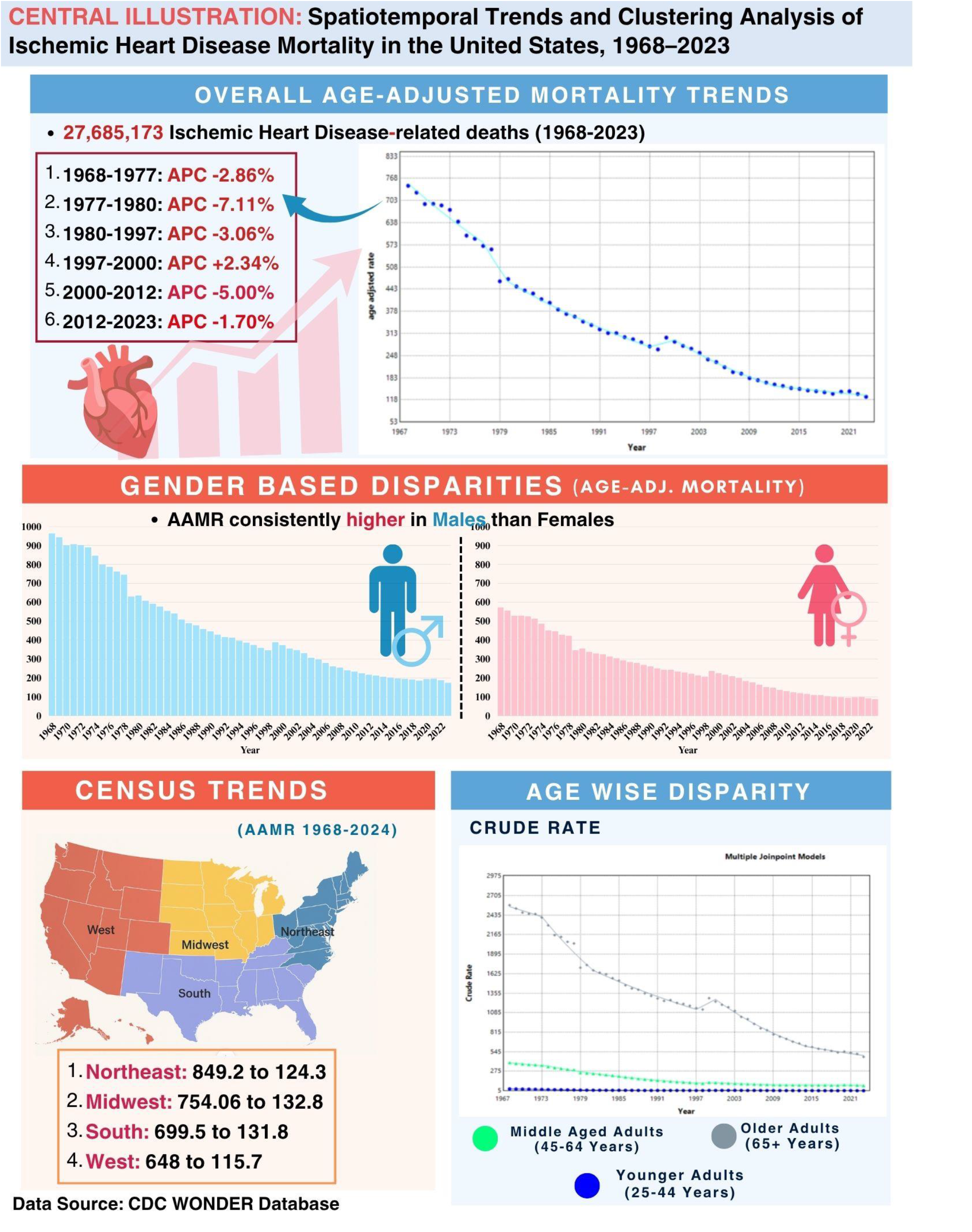
Central Illustration.

**Figure 2:**
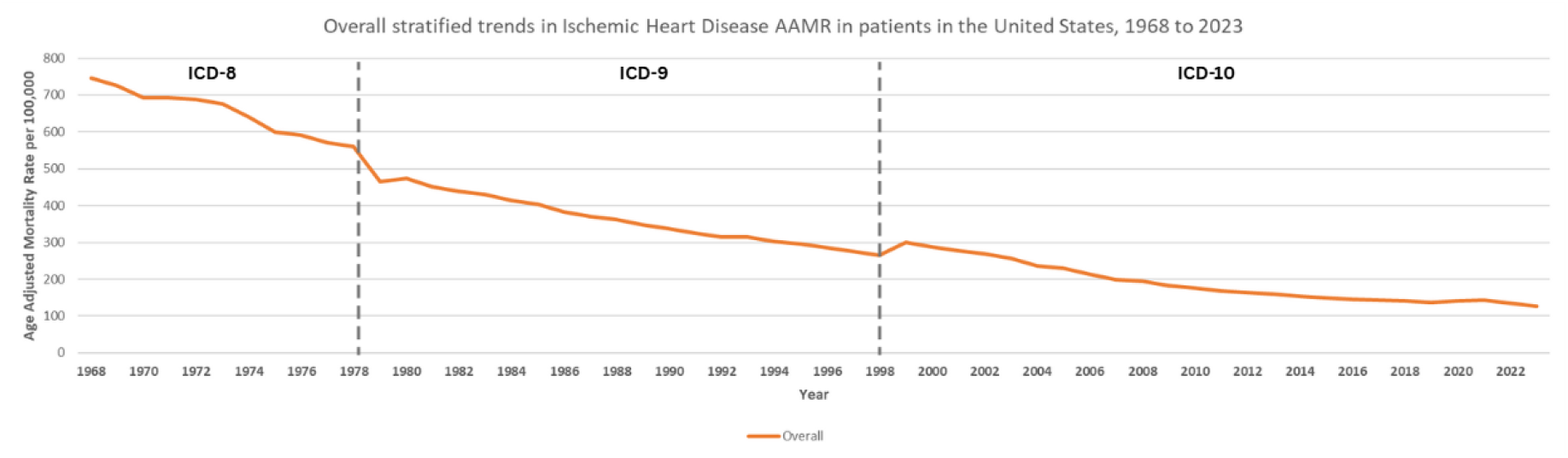
Overall stratified trends in Ischemic Heart Disease AAMR in patients in the United States, from 1968 to 2023.

Between 1968 and 1977, mortality fell moderately (APC –2.86; 95% CI, –3.28 to –2.44; *p*<0.001), followed by a sharper drop through 1980 (APC –7.11; 95% CI, –11.51 to –2.48; *p*=0.004). The downward trend continued consistently from 1980 to 1997 (APC –3.06; 95% CI, –3.26 to –2.87; p<0.001).

A brief, nonsignificant rise occurred between 1997 and 2000 (APC 2.34; 95% CI, –2.85 to 7.80; *p*=0.37), after which mortality again declined sharply from 2000 to 2012 (APC –5.00; 95% CI, – 5.36 to –4.64; p<0.001). From 2012 onward, the rate continued to drop but at a slower pace (APC –1.70; 95% CI, –2.10 to –1.29; p<0.001).

Overall, across the 55-year period, mortality fell by an average of 3.1% per year (AAPC –3.12; 95% CI, –3.51 to –2.73; p<0.001), reflecting a sustained, long-term improvement in outcomes.

### Gender Trends

From 1968 to 2023, the AAMR was higher among males than females (452.99 vs. 260.08) as shown in **(Supplementary Table 2 & 3, Fig 3)**.

**Figure 3:**
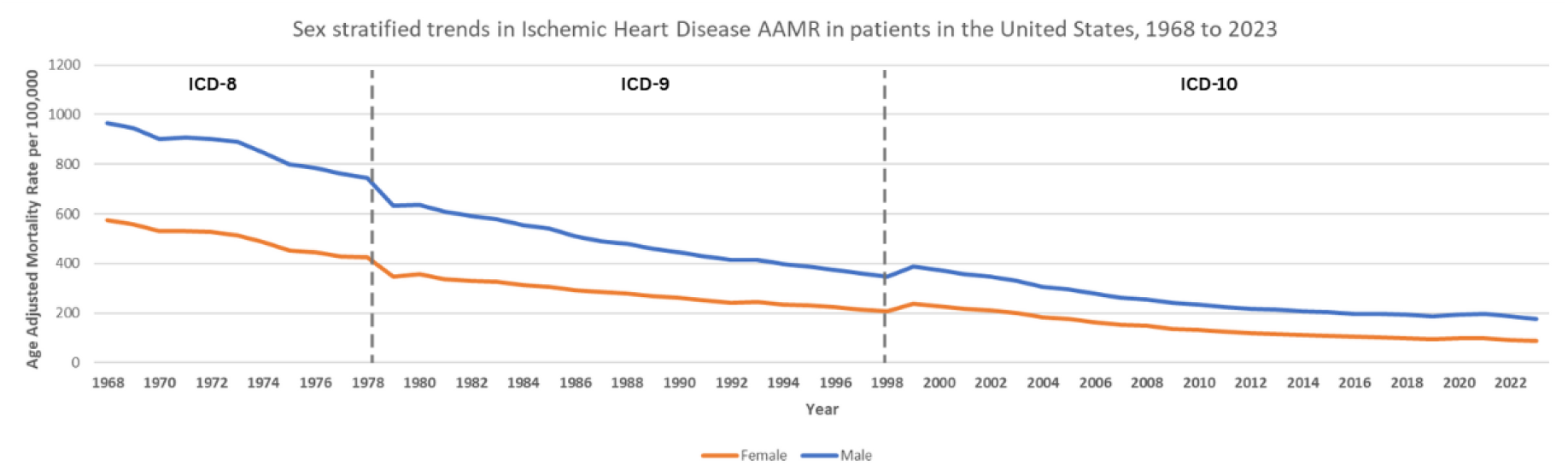
Sex stratified trends in Ischemic Heart Disease AAMR in patients in the United States, from 1968 to 2023.

Among males, mortality declined significantly across multiple periods. Between 1968 and 1977, the AAMR decreased at APC of –2.50 (95% CI: –2.93 to –2.07; p<0.001), followed by a steeper decline from 1977 to 1980 (APC –6.82; 95% CI: –11.43 to –1.96; p=0.008). A persistent reduction was then observed from 1980 to 1997 (APC –3.25; 95% CI: –3.45 to –3.05; p<0.001).

Between 1997 and 2000, no significant change occurred (APC 1.71; 95% CI: –3.86 to 7.60; p=0.55). The downward trend resumed sharply between 2000 and 2012 (APC –4.66; 95% CI: – 5.03 to –4.28; p<0.001), followed by a more gradual decline from 2012 to 2023 (APC –1.36; 95% CI: –1.76 to –0.95; p<0.001). Over the entire 1968–2023 period, the AAPC was –3.00 (95% CI: –3.00 to –3.00; p<0.001), reflecting a sustained long-term reduction in male mortality.

Among females, mortality followed a similar overall declining trajectory. From 1968 to 1976, the AAMR decreased at an APC of –2.76 (95% CI: –3.27 to –2.25; p<0.001), followed by an accelerated decline from 1976 to 1979 (APC –7.44; 95% CI: –11.69 to –2.98; p=0.002). Between 1979 and 1997, a consistent decrease persisted (APC –2.92; 95% CI: –3.09 to –2.75; p<0.001). A brief nonsignificant increase occurred from 1997 to 2000 (APC 2.94; 95% CI: –2.14 to 8.29; p=0.25), followed by a sharp decline between 2000 and 2012 (APC –5.55; 95% CI: –5.90 to – 5.19; p<0.001). From 2012 to 2023, the rate continued to decline modestly (APC –2.39; 95% CI: –2.81 to –1.96; p<0.001). Overall, from 1968 to 2023, female mortality decreased significantly with an AAPC of –3.00% (95% CI: –4.00 to –3.00; p<0.001).

### Race Trends

From 1968 to 2023, White individuals exhibited a slightly lower mean age-adjusted mortality rate (345.38) compared with Black or African American individuals (346.75). However, the temporal patterns of decline differed notably between the two groups **(Supplementary Table 2 & 4, Figure 4)**. Among Black or African American populations, mortality dropped steadily from 1968 to 1977 (APC –2.88; 95% CI, –3.47 to –2.29; p<0.001), followed by a sharper decline through 1980 (APC –9.79; 95% CI, –15.95 to –3.18; p=0.005). From 1980 to 1997, the decline continued at a slower yet consistent pace (APC –2.05; 95% CI, –2.33 to –1.77; p<0.001).

**Figure 4:**
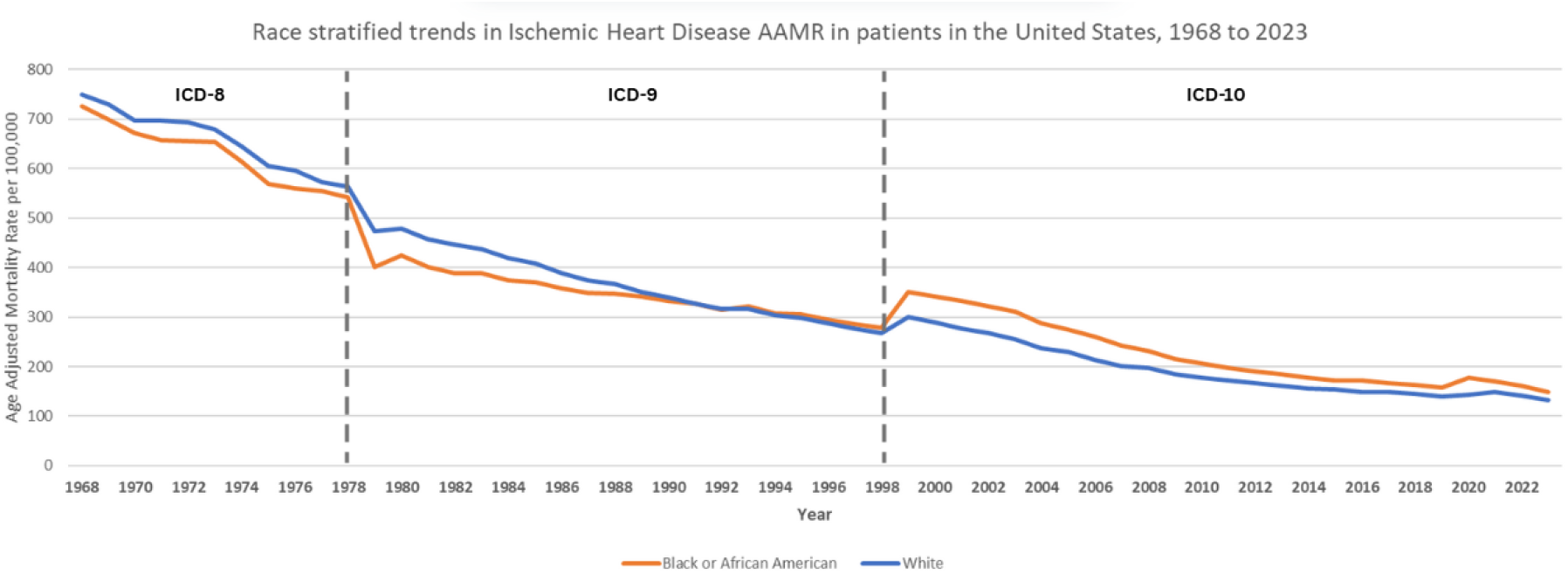
Race stratified trends in Ischemic Heart Disease AAMR in patients in the United States, from 1968 to 2023.

A brief, nonsignificant rebound appeared between 1997 and 2000 (APC 7.13; 95% CI, –0.10 to 14.87; p=0.053), but mortality resumed a marked decline from 2000 to 2013 (APC –5.09; 95% CI, –5.49 to –4.69; p<0.001). In the last decade, 2013–2023, progress slowed but remained significant (APC –1.26; 95% CI, –1.84 to –0.68; *p*<0.001).

Overall, across the full 56-year period, mortality decreased by an average of 2.7% per year (AAPC –2.73; 95% CI, –3.27 to –2.19; p<0.001), underscoring sustained long-term improvement.

Among White populations, mortality trends showed a similar but slightly steeper overall improvement. Rates fell moderately from 1968–1976 (APC –2.56; 95% CI –3.03 to –2.09; p<0.001), followed by an accelerated decline between 1976–1979 (APC –6.91; 95% CI –10.94 to –2.71; p=0.002). A long phase of gradual improvement occurred from 1979–1997 (APC – 3.20; 95% CI –3.37 to –3.04; p<0.001), interrupted briefly by a nonsignificant increase between 1997–2000 (APC 2.18; 95% CI –2.87 to 7.49; p=0.39). Between 2000 and 2012, mortality again decreased sharply (APC –4.83; 95% CI –5.19 to –4.48; p<0.001), followed by a modest but significant decline from 2012–2023 (APC –1.51; 95% CI –1.92 to –1.09; p<0.001). Over the full period, the AAPC confirmed a sustained reduction (AAPC –3.05; 95% CI –3.42 to –2.68; p<0.001). Although both groups achieved major reductions since 1968, the decline in mortality was slightly steeper among White individuals, leading to a gradual narrowing but persistent racial gap through 2023.

### Census Region Trends

Throughout the study period (1968-2023), the Northeast had the highest overall average annual mortality rate (379.61), followed by the Midwest (351.31), the South (329.15), and the West (303.15**). (Supplementary Table 2 & 5, Figure 5)**.

**Figure 5:**
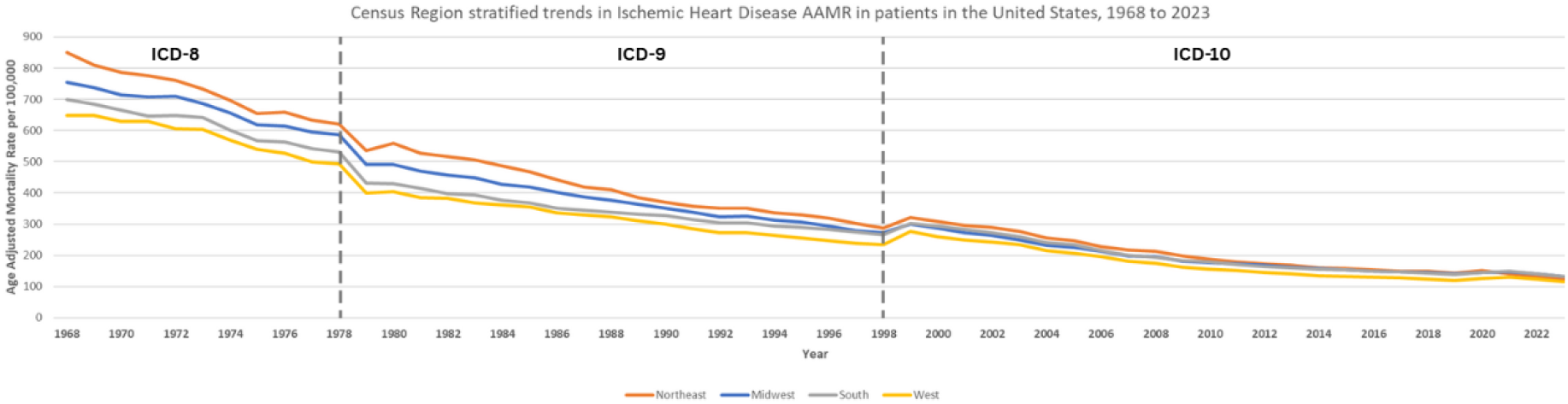
Census Region stratified trends in Ischemic Heart Disease in patients in the United States, from 1968 to 2023.

In the Northeast, mortality decreased significantly from 1968 to 1997 (APC −3.60; 95% CI −3.70 to −3.50; p<0.001). A non-significant increase was observed between 1997 and 2000 (APC 1.97; 95% CI −5.32 to 9.83; p=0.60), followed by a very rapid decline from 2000 to 2012 (APC −4.97; 95% CI −5.49 to −4.46; p<0.001). The rate of decline slowed from 2012 to 2023 but remained significant (APC −2.39; 95% CI −3.01 to −1.77; p<0.001). Across the entire 55-year interval, the AAMR declined significantly (AAPC −3.37; 95% CI −3.78 to −2.95; p<0.001).

In the Midwest, an initial period of decline occurred from 1968 to 1977 (APC −2.56; 95% CI −2.95 to −2.18; p<0.001), which accelerated dramatically between 1977 and 1980 (APC −6.96; 95% CI −11.10 to −2.63; p=0.003). A steady, significant decline continued from 1980 to 1997 (APC −3.14; 95% CI −3.33 to −2.96; p<0.001). After a period of non-significant change from 1997 to 2000 (APC 0.69; 95% CI −4.50 to 6.17; p=0.79), a sharp decline resumed from 2000 to 2011 (APC −4.87; 95% CI −5.29 to −4.44; p<0.001). The final segment from 2011 to 2023 saw a more modest but significant decline (APC −1.69; 95% CI −2.07 to −1.31; p<0.001). Overall, the AAMR declined significantly (AAPC −3.09; 95% CI −3.47 to −2.70; p<0.001).

In the South, mortality fell from 1968 to 1977 (APC −2.70; 95% CI −3.19 to −2.21; p<0.001), with a period of accelerated decline from 1977 to 1980 (APC −8.87; 95% CI −13.79 to −3.67; p=0.002). A steady decrease continued from 1980 to 1997 (APC −2.50; 95% CI −2.72 to −2.29; p<0.001). A non-significant increase was noted between 1997 and 2000 (APC 3.20; 95% CI −2.34 to 9.05; p=0.25), followed by the steepest decline in the region from 2000 to 2012 (APC −5.07; 95% CI −5.44 to −4.70; p<0.001). The pace of decline slowed from 2012 to 2023 but remained significant (APC −1.45; 95% CI −1.86 to −1.04; p<0.001). Across the full interval, the South demonstrated a significant decline (AAPC −2.95; 95% CI −3.38 to −2.52; p<0.001).

In the West, an initial decline occurred from 1968 to 1976 (APC −2.50; 95% CI −3.08 to −1.91; p<0.001), which accelerated sharply from 1976 to 1979 (APC −8.13; 95% CI −12.98 to −3.00; p=0.003). A prolonged period of significant decline followed from 1979 to 1997 (APC −3.09; 95% CI −3.29 to −2.90; p<0.001). A non-significant increase was observed between 1997 and 2000 (APC 3.66; 95% CI −1.86 to 9.49; p=0.19), before a very rapid decline from 2000 to 2013 (APC −5.03; 95% CI −5.35 to −4.71; p<0.001). The final segment from 2013 to 2023 showed a slower but significant rate of decrease (APC −1.30; 95% CI −1.76 to −0.83; p<0.001). Over the entire study period, the West showed a significant decline in AAMRs (AAPC −3.07; 95% CI −3.49 to −2.65; p<0.001).

### Age groups Trends

Across 1968–2023, crude mortality (CMR) differed markedly by age. Older adults (≥65 years) had the highest mean CM (1312.66; 95% CI: 1308.70–1316.63), followed by middle-aged adults (45–64 years) (171.67; 95% CI: 170.60–172.74). Younger adults (25–44 years) had the lowest rate (12.18; 95% CI: 11.93–12.43). **(Supplementary Table 2 & 6, Figure 6)**.

**Figure 6:**
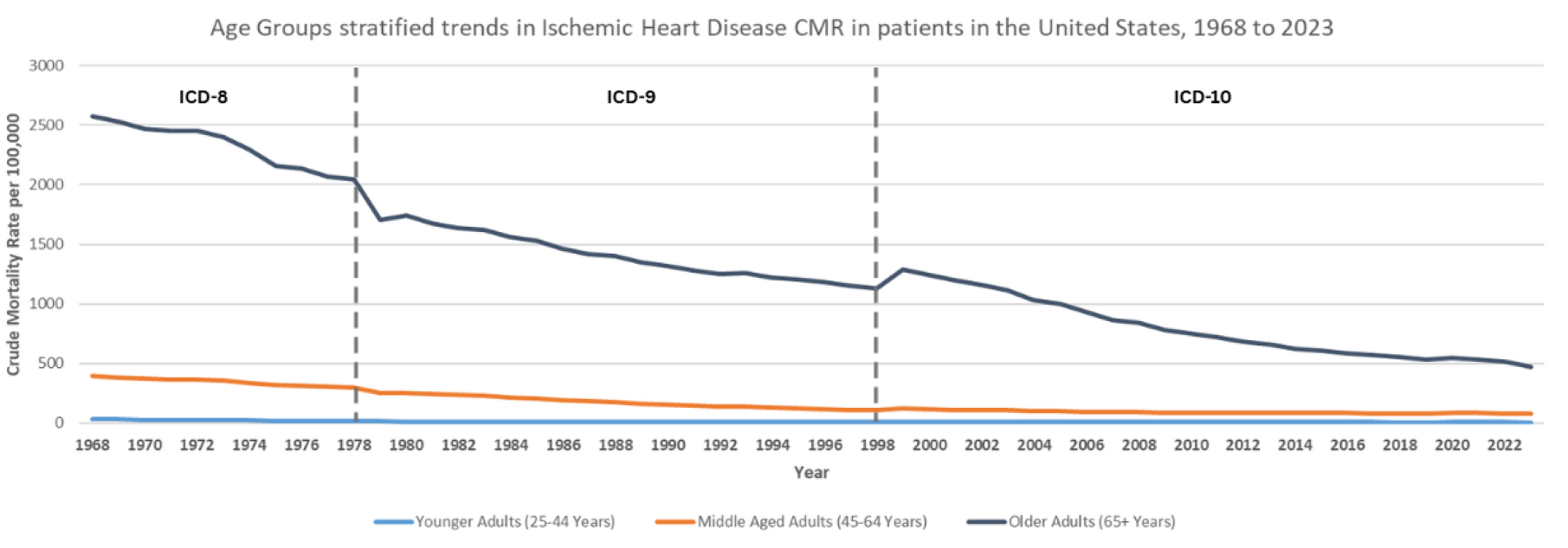
**Age-**groups stratified in the Ischemic Heart Disease AAMR **in patients in the United States, from 1968 to 2023**. *Spatiotemporal Clustering (1979-2023):*

Among younger adults (25–44 years), CMRs showed several fluctuations across the study period. After early declines in the 1970s and 1980s (e.g., 1968–1971 APC −4.02; 95% CI −6.74 to −1.23; p=0.006 and 1971–1980 APC −7.30; 95% CI −7.97 to −6.63; p<0.001), trends flattened in the 1990s (1990–1997 APC −0.83; 95% CI −2.14 to 0.49; p=0.21). A significant rise occurred from 1997–2002 (APC 3.17; 95% CI 0.71 to 5.70; p=0.013), followed by a sustained decline from 2002–2018 (APC −3.35; 95% CI −3.69 to −3.00; p<0.001). Rates trended upward but non-significantly during 2018–2021 (APC +5.58; 95% CI −3.47 to 15.49; p=0.23) and then declined again in 2021–2023 (APC −7.30; 95% CI −15.40 to 1.57; p=0.10). Over the full 1968– 2023 interval the average annual percent change (AAPC) was −2.91 (95% CI −3.57 to −2.26; p<0.001), indicating a significant long-term reduction in CMR for this age group despite short-term fluctuations.

For middle-aged adults (45–64 years), mortality fell steeply in the late 20th century. Early modest declines were followed by a pronounced reduction from 1974–1997 (APC −4.79; 95% CI −4.94 to −4.64; p<0.001). A nonsignificant uptick occurred in 1997–2000 (APC 1.35; 95% CI −6.26 to 9.57; p=0.73), but a renewed decline was evident from 2000–2010 (APC −3.21; 95% CI −3.88 to −2.54; p<0.001). From 2010–2023 the trend leveled (APC −0.27; 95% CI −0.67 to +0.14; p=0.19). Across the entire study period the AAPC was −2.83 (95% CI −3.27 to −2.39; p<0.001), reflecting a sustained long-term decrease in IHD crude mortality in middle-aged adults.

In older adults (≥65 years), declines were large and consistent over most decades. After a weak early change (1968–1973 APC −1.10; p=0.09), a sharp reduction occurred from 1973–1981 (APC −4.51; 95% CI −5.26 to −3.74; p<0.001) and a continued decline through 1981–1997 (APC −2.41; 95% CI −2.67 to −2.15; p<0.001). A transient, nonsignificant rise was observed in 1997–2000 (APC 3.95; 95% CI −2.45 to 10.76; p=0.23), followed by a marked fall from 2000– 2014 (APC −4.99; 95% CI −5.34 to −4.65; p<0.001) and a continued but slower decline from 2014–2023 (APC −2.49; 95% CI −3.16 to −1.81; p<0.001). The overall AAPC for older adults was −2.94 (95% CI −3.33 to −2.55; p<0.001), indicating a sustained, statistically significant long-term reduction in crude IHD mortality for this group.

**(Supplementary Table 7, Figure 7 & 8)**.

**Figure 7:**
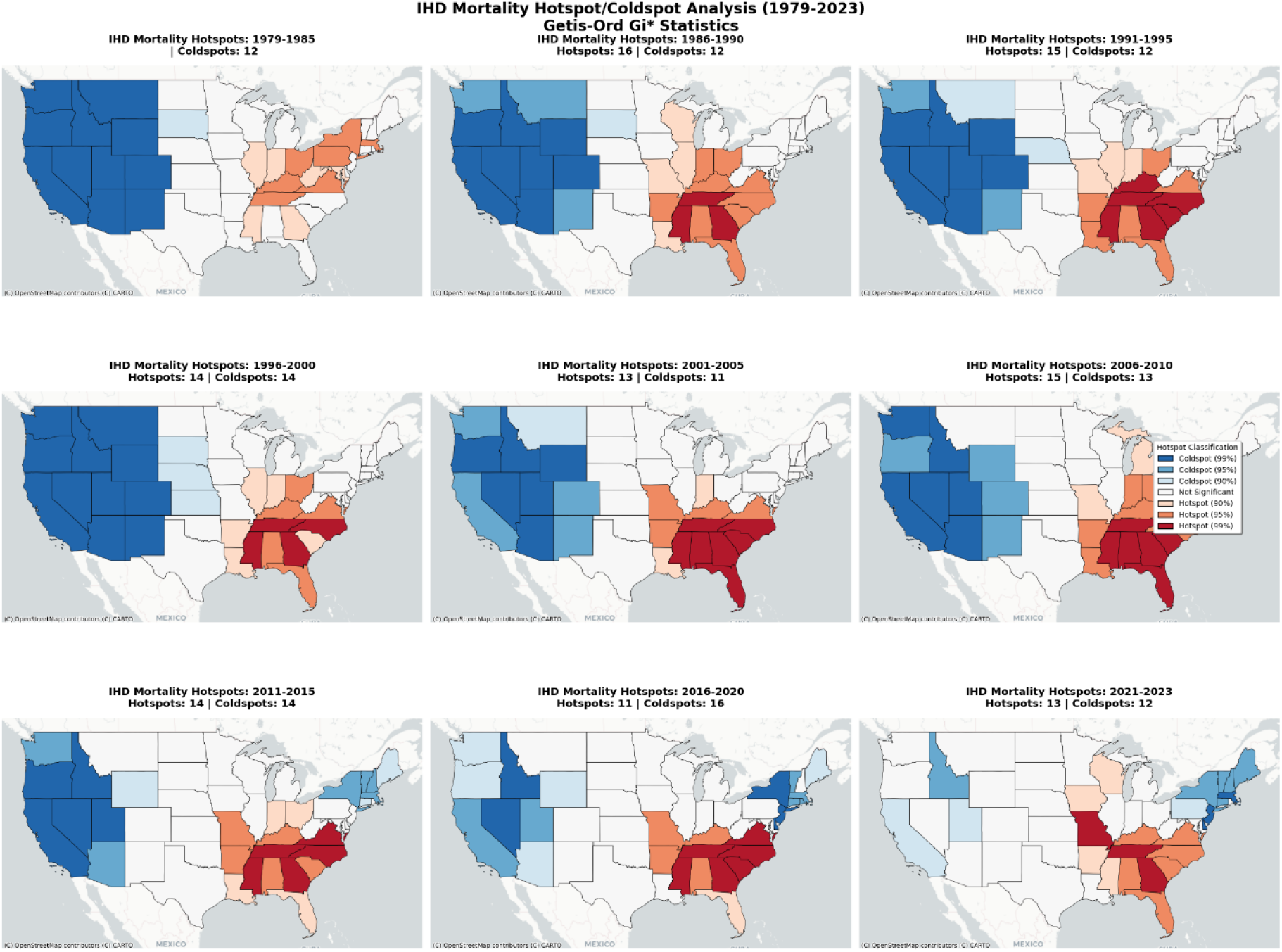
Spatiotemporal clustering of ischemic heart disease (IHD) mortality in the United States, 1979–2023. (reason for 1968-1978 period exclusion is described in method section) Each panel represents a five-year interval (except the first and last) showing statistically significant hotspots (red) and coldspots (blue) based on Getis-Ord Gi* statistics. Hotspots indicate clusters of states with significantly higher IHD mortality rates, while coldspots represent regions with significantly lower mortality.

**Figure 8:**
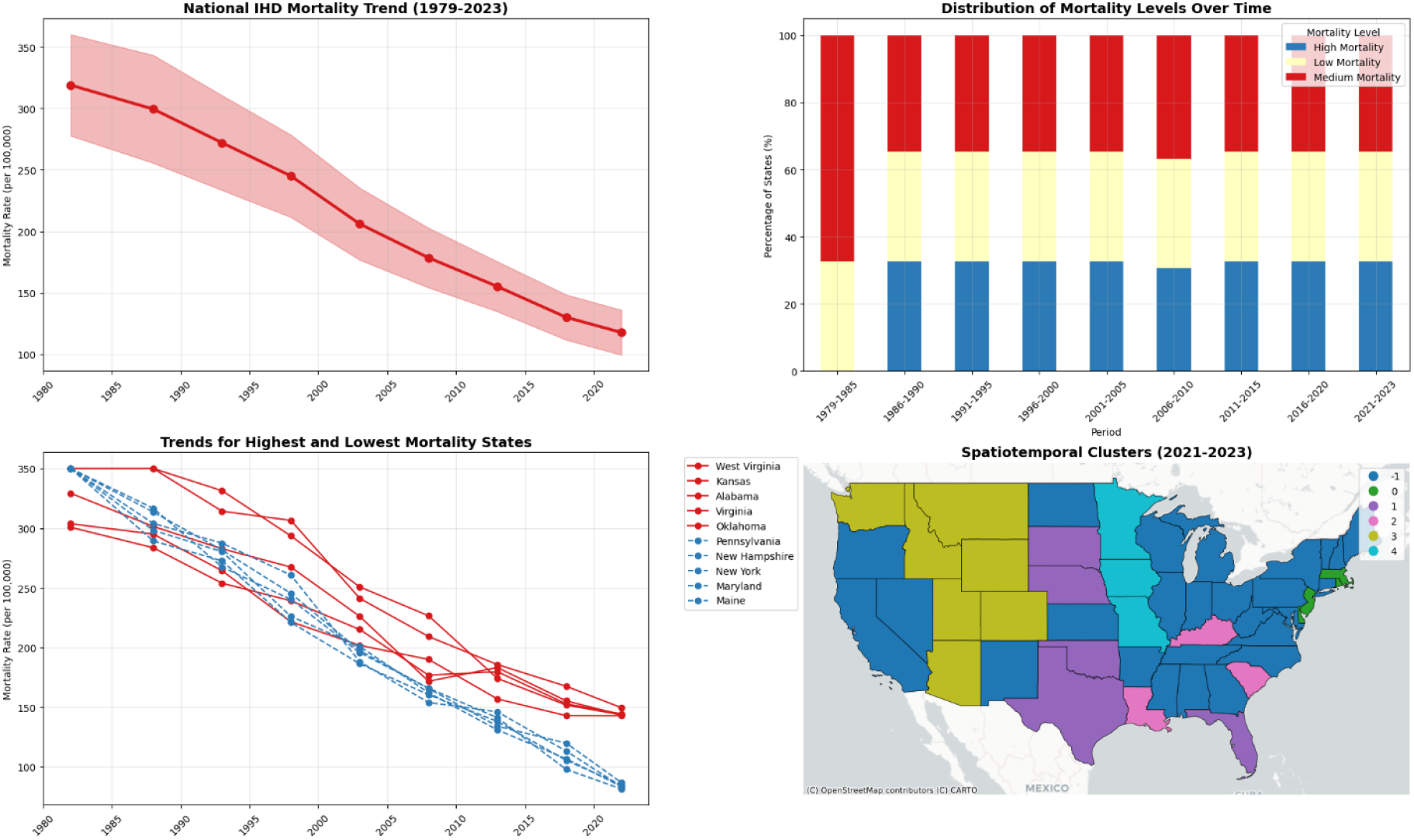
Temporal and spatial trends in ischemic heart disease (IHD) mortality in the United States, 1979–2023. **(A) National IHD Mortality Trend:** The age-adjusted mortality rates (AAMR) declining across the study period. Shaded areas indicate 95% confidence intervals. **(B) Distribution of Mortality Levels Over Time:** The proportion of high-mortality states increased from 0% in 1979–1985 to 16 states (32%) by 2021–2023, while the share of low-mortality states remained constant. **(C) Trends for Highest and Lowest Mortality States:** Persistently high rates were observed in West Virginia, Kansas, and Alabama, whereas states such as Pennsylvania, New York, and New Hampshire maintained lower mortality trajectories throughout the study period. **(D) Spatiotemporal Clusters (2021–2023):** Cluster analysis identified distinct spatial groupings, with persistent hotspots in the Southeast and coldspots in the Western and Mountain regions, underscoring an enduring east–west mortality divide despite national improvements.

Clear geographic clustering of IHD mortality emerged over time. In the early period (1979– 1985), spatial distribution was diffuse with no significant high-mortality clusters. By 2021–2023, strong clusters formed across the Southeast and central regions.

High-mortality clusters were consistently seen in Alabama, Kentucky, Tennessee, Georgia, and West Virginia. These states showed stable spatial persistence, indicating long-standing regional risk concentration. In contrast, low-mortality clusters remained concentrated in the western region, including Washington, Oregon, Utah, and Colorado.

Hotspot intensity increased between 2000 and 2010, then stabilized. Coldspots in the Mountain and Pacific regions persisted with minimal temporal variation. This spatial polarization highlights a widening geographic divide in IHD mortality despite overall national declines.

## Discussion

The nationwide evaluation of ischemic heart disease (IHD) mortality from 1968 to 2023 has shown a notable and steady decrease in mortality rates in the United States across all demographics, geographies, and age cohorts. Between 1977 and 2012, the greatest declines were recorded, owing to the cumulative effects of the advances in the management of acute cardiac care, the implementation of evidence-based practices, and the active management of hypertension, hyperlipidemia, smoking, and smoking cessation. The slow rate of decline after 2012 suggests a plateauing of the benefits of interventions and the burdens of skyrocketing obesity and diabetes, as well as health inequity. While all age groups experienced these inequities and health burdens, the greatest absolute burden of mortality for IHD was among the 65 and older age group, thus the broad public health advances. The excess mortality burden was seen among men, and although both men and women experienced the inequity of improvements. This suggests the need to address the excess mortality burden on men. The improvements for the black and the white populations underline the need for interventions that prevent inequities. The South demonstrated the greatest excess mortality and the lowest rate of improvement. There were improvements across all age groups, which was a sign of broad public health progress, even though relative enhancements were also observed across all age groups, despite progress on healthcare and service equity.

According to the INTERHEART Study, the risk factors that are responsible for more than 90% of the risk of heart conditions like IHD and acute MI include smoking, diabetes, obesity, poor diet, inactivity, excessive alcohol, hypertension, dyslipidemia, and psychosocial stress. The declines in some of these factors accounted for 44% of the decline in IHD mortality in the U.S., but the prevalence of diabetes and obesity holds steady, and even increases. Optimal risk profiles translate to significantly lower lifetime risk of MI, stroke, and cardiovascular death, yet fewer than 2% of U.S. adults meet ideal cardiovascular health.^**17**^

In both sexes, the decline was significant in the long term, but mortality in males was consistently higher than that of females. The decline in mortality was steepest in the period 1977–2012, with modest changes after 2012 in both sexes. In the acute coronary syndrome (ACS) and during percutaneous coronary intervention referrals, women were noted to have less coronary atherosclerosis, which is contrary to expectations.^**18**^ Even with a greater burden of comorbidity, as shown in the PROSPECT study, women often present with non-obstructive or less severe CAD.^**18,19**^ It has been suggested that the endothelial and microvascular dysfunction partially explains women’s ischemic symptoms even with minimal obstruction.^**19,20**^ Han et al. showed that men had longer arterial segments with endothelial dysfunction, and women had lower coronary flow reserve, consistent with microvascular impairment.^**19**^ The WISE study linked impaired coronary flow reserve with adverse outcomes in women without obstructive CAD.^**20**^ Experimental models show that female hearts experience reduced apoptosis and infarct size during ischemia–reperfusion, likely due to estrogen-mediated cardioprotection.^**21,22**^ Clinically, women have a higher risk of heart failure (HF) complicating acute MI,^**23,24**^ though men exhibit a greater risk of sudden cardiac death and ventricular arrhythmias post-MI.^**25,26**^ These sex-specific differences highlight the interplay between hormonal, microvascular, and structural mechanisms influencing outcomes in ischemic heart disease.

Both White and Black populations experienced substantial mortality reductions since 1968, though declines were slightly steeper among Whites. Racial gaps narrowed gradually but persisted through 2023. While trivial risk factors like hypertension, diabetes, obesity, and smoking contribute to elevated CAD and HF risk, these do not fully explain the disproportionate burden in minority populations.^**27**^ Disparities such as lower socioeconomic status, lack of access to healthy foods, safe neighbourhoods, and quality medical care, and lack of access to care contribute to risk.^**28**^ Children and adolescents from communities with racial deprivations, especially Black children, exhibit signs of hypertension at younger ages and carry it to adulthood.^**29-31**^

Chronic psychosocial adversities such as racial discrimination, unsteady employment, and living in high-crime areas contribute to cardiovascular risk via inflammation and the neuroendocrine system and dysregulation.^**32,33**^ Information from the GWTG-HF registry and others has documented that Black and Hispanic patients receive therapies such as β-blockers, ICDs, and cardiac rehabilitation less frequently than White patients and are less likely to receive therapies.^**34**^ Asian patients with HFpEF have the highest risk of in-hospital mortality.^**35**^ Disparities in outcomes, even after controlling for risk factors, point to the importance of factors beyond the health care system. Social determinants of health, systemic inequities, and possible biological predisposition reflect the complexity of the problem. Effective culturally based public health approaches, increased care access, and interventions that are pre-emptive and equitable are necessary to reduce IHD mortality in high-risk and vulnerable racial and ethnic populations.

All U.S. regions demonstrated significant long-term declines, with the steepest improvements in the Northeast and West. The South consistently had the highest mortality and also the slowest rate of decline. These disparities likely reflect regional differences in socioeconomic status, healthcare access, and the prevalence of chronic comorbid conditions, such as obesity and hypertension. Geographic variations in AAMR for IHD likely stem from differences in healthcare access, socioeconomic status, and prevalence of modifiable risk factors like obesity, smoking, and hypertension.

The mortality rates have declined significantly in all age groups, with the adults aged 65 and above having the largest absolute burden. The younger and middle-aged adults have also demonstrated prolonged reductions in mortality rates; however, they have experienced some short-term fluctuations. These unstable trends in the young group of patients can be attributed to the findings suggesting that they have a significantly higher rise in rates of dyslipidemia and obesity as compared to the older patients, which is in line with the studies that reported dyslipidemia in 2.5–97.3% of very young CAD cases.^**36,37**^ The prevalence of hypertension in older patients has been reported to vary between 8.8% and 80.0%, which is higher than that of young adults.^**38**^ Moreover, the use of smokeless tobacco has been singled out as one of the newer contributing risk factors, with the frequencies among acute MI patients ranging from 0.3% to 60.6%.^**39**^ In contrast, diabetes mellitus and hypertension were significantly more common in the elderly group, which indicates that these conditions are the main contributors to the occurrence of first-time AMI in this group of patients.

The present analysis demonstrates substantial progress across the whole nation in reducing IHD mortality over the past five decades, coinciding with improvements in risk factor management, acute cardiovascular care, and secondary prevention. Mean mortality declined from more than 600 per 100,000 population in 1968–1978 to fewer than 170 per 100,000 population in 2011– 2020, reflecting major advances in public health and clinical interventions. However, the magnitude of reduction was not uniform across the United States. Hotspot counties consistently exhibited markedly higher mortality than in coldspots in each period, indicating that regional inequities in cardiovascular outcomes have not only persisted but have become more geographically established.

A distinct spatiotemporal redistribution of risk was evident. Mid-century hotspots in the industrial Midwest and central United States gradually shifted toward the Southeast and lower Mississippi Valley, areas characterised by elevated burdens of poverty, conditions like stroke,^**40**^ hypertension, diabetes,^**41**^ heart failure,^**42**^ and structural barriers to preventive and speciality care.^**43**^ Coldspots grew more concentrated in the Mountain West and New England, regions with stronger healthcare access and healthier population profiles.^**44**^ The persistence and intensification of this geographic divide underscore that continued reductions in national IHD mortality will require targeted prevention strategies, investment in healthcare infrastructure, and efforts to mitigate the social and structural determinants that sustain regional vulnerability.^**44**^

Implementation frameworks such as the Practical, Robust Implementation and Sustainability Model (PRISM) facilitate policy translation by systematically linking evidence to real-world practice. They help identify contextual factors influencing adoption, scalability, and sustainability, thereby enhancing the effectiveness and equity of implemented interventions. For example in the context of cardio-oncology, the integration of frameworks like PRISM alongside RE-AIM and CFIR has been highlighted as enabling systematic attention to reach, adoption, implementation, and maintenance of interventions.^**45**^

New studies should aim at clarifying the concepts of the complex factors that are responsible for slow decline of IHD mortality. The main focus should be on the investigation of the onset of cardiometabolic risk factors, such as obesity, diabetes, and lifestyle-related behaviours and that could harm the progress made so far. Improved monitoring systems that combine clinical, socioeconomic, and geographic data would be useful in the identification of vulnerable subpopulations and areas at high risk with inefficient preventive measures and inadequate healthcare facilities. Moreover, studies should evaluate the effects of healthcare access, preventive cardiology programs, and regional policy initiatives on disparities. Utilisation of electronic health records and genomic data may pave the way to develop precision prevention strategies that address vulnerabilities related to all the demographic subgroups. Constant efforts related to cardiovascular prevention, acute care management, fair healthcare provision, and health education will be essential to curb the death rates in the future.

## Limitations

This study relied on death certificate data, which are subject to potential misclassification and diagnostic inconsistencies, particularly in earlier years. Changes in International Classification of Diseases (ICD) coding over time may have influenced trend comparability. When utilising collective data, we lose the ability to adjust for factors such as individual socioeconomic standing, comorbidities, and access to healthcare. Population migration patterns and shifts in age structure over the study period may also have affected observed trends, as both can alter the demographic composition and disease burden of populations. In addition, the influence of preventive measures, advancements in treatment, and changes in behaviour concerning risks could be evaluated in this analysis. Finally, the geographical and ethnic categories employed may obscure diversity and changes in population structure within the studied groups, a factor that has altered over the last fifty years.

## Conclusion

Over the last fifty years, the US has seen the most profound and positive changes in the management, prevention, and post-acute care of cardiovascular conditions, particularly in the case of IHD, which resulted in the IHD mortality rates within the country declining. This post-acute care has not only improved in the management of the IHD, but also in the long-term care of the cardiovascular conditions and the cumulative benefits of treatment. These changes have been comprehensive as they have been observed in all sexes, races, and ages. Nevertheless, IHD management has inequities, particularly concerning racial and geographical access disparities.

## Abbreviations

AAMR: Age-Adjusted Mortality Rate
AAPC: Average Annual Percent Change
APC: Annual Percent Change
ICD: International Classification of Diseases
IHD: Ischemic Heart Disease

## Declarations

## Acknowledgements

None

## Ethical Approval

Not applicable

## Data availability statement

All data supporting the findings of this report are available within the article and can be accessed from https://wonder.cdc.gov/deaths-by-underlying-cause.html

## STROBE Statement

Checklist of items that should be included in reports of *cross-sectional studies*

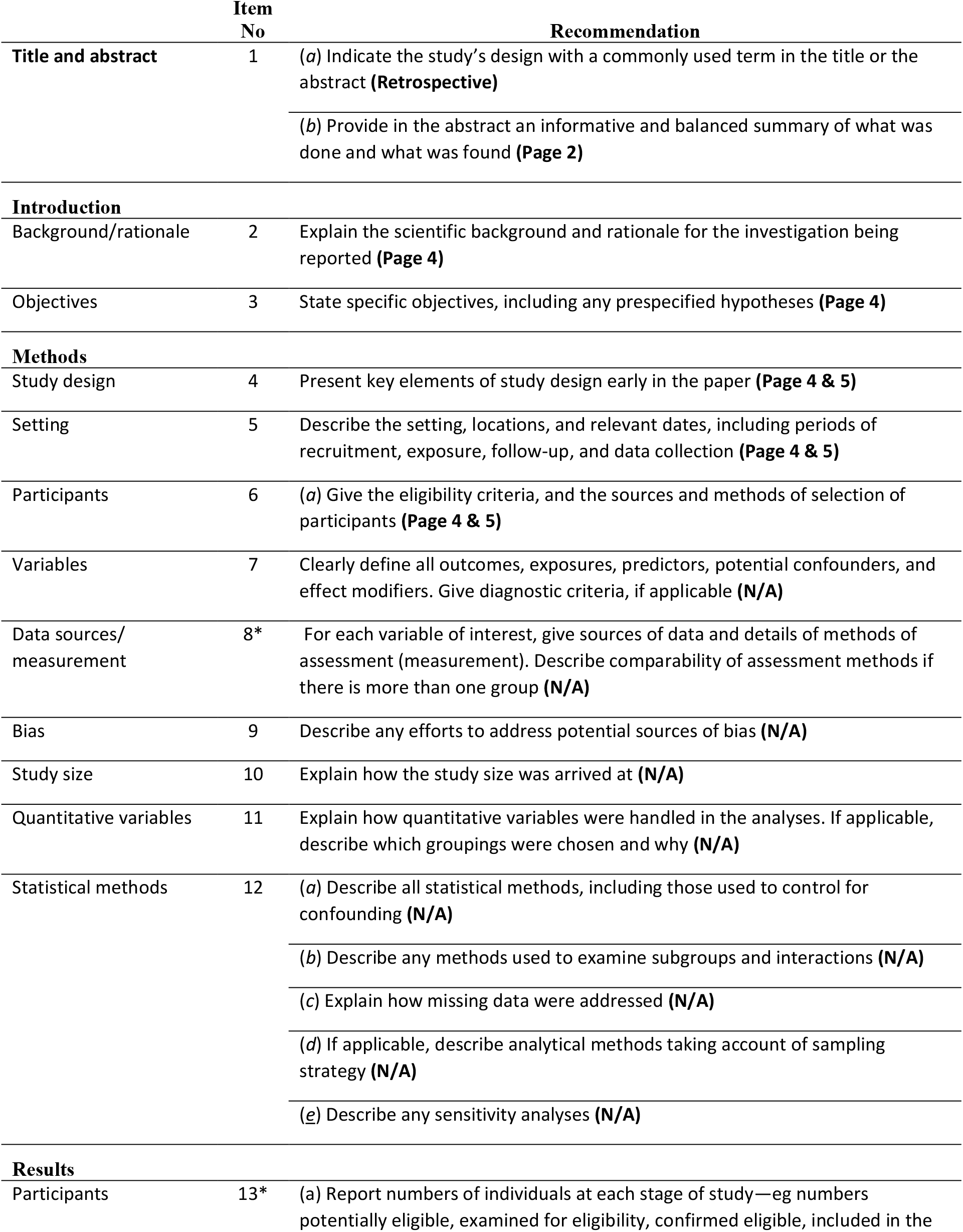
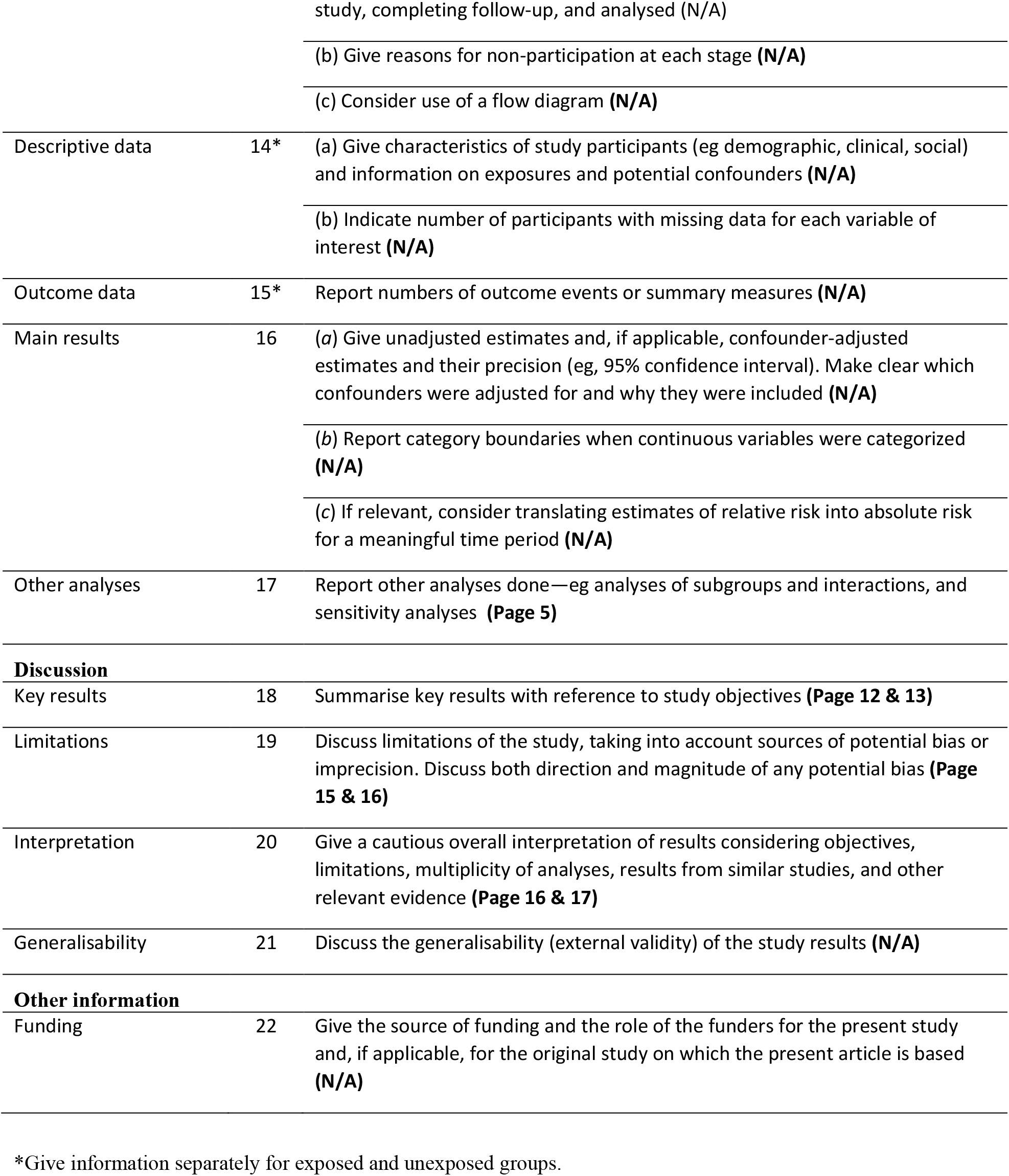

